# Molecular counting enables accurate and precise quantification of methylated ctDNA for tumor-naive cancer therapy response monitoring

**DOI:** 10.1101/2023.05.31.23290555

**Authors:** Patrick Peiyong Ye, Robb Andrew Viens, Katherine Elise Shelburne, Sydne Scot Langpap, Xavier Scott Bower, Wen Zhou, Jan Christian Wignall, Joyce Jiawei Zhu, Brian D Woodward, Hatim Husain, David S Tsao, Oguzhan Atay

## Abstract

Personalized cancer treatment can significantly extend survival and improve quality of life for many patients, but accurate and real-time therapy response monitoring remains challenging. To overcome logistical and technical challenges associated with using imaging scans or assays that track the variant allele fraction (VAF) of somatic mutations in circulating tumor DNA (ctDNA) for response monitoring, we developed a tumor-naive liquid biopsy assay that leverages quantitative counting template (QCT) technology to accurately and precisely quantify methylated ctDNA (Northstar Response^TM^). Northstar Response^TM^ achieves <10% coefficient of variation at 1% tumor fraction, which is 2x lower than VAF-based response monitoring approaches. The assay accurately distinguishes 0.25% absolute changes in contrived tumor fraction (AUC > 0.94) and performs well in 12 solid tumor types. Preliminary clinical results from patients with lung, colorectal, or pancreatic cancer demonstrate that Northstar Response^TM^ detects changes in ctDNA methylation that correlate with clinical outcomes. As a novel tool for therapy response monitoring, the assay’s serial measurements of ctDNA methylation can be precise, reflect clinical outcomes, and have potential to inform clinical decision making for cancer treatment.

## Introduction

While novel cancer treatments continue to be developed at an unprecedented pace, assessing whether a cancer treatment is effective for a particular patient remains cumbersome and relatively subjective and qualitative. Imaging is the current standard for measuring the state of a patient’s cancer; however, imaging faces limitations in accuracy and precision for therapy response monitoring. Specifically, a patient’s response to therapy may not be accurately reflected on imaging due to tumoral heterogeneity^1^, pseudoprogression in patients receiving immunotherapy^2^, scar tissue surrounding the tumor, or low contrast in certain tissues like bone and the peritoneum^3–5^. These measurements may also be imprecise due to the subjectivity involved with measuring radiographic images^1^. In addition, the relative infrequency of imaging, with scans often several months apart, lengthens the lead time for assessing response to treatment and identifying when a patient is progressing on a particular therapy. Increasing the frequency of imaging can be infeasible due to access, logistics of traveling to imaging centers, and cost. Protein biomarkers have some but limited value for monitoring, with varying utility across cancer types^6^. There is a need to more accurately, precisely, and frequently assess the extent to which a patient’s cancer is responding to treatment to inform clinical decision making.

Non-invasive liquid biopsies that assay cell-free DNA (cfDNA) have been used to quantify the levels of circulating tumor DNA (ctDNA) from a blood sample. Several studies have found changes in ctDNA levels to be predictive of progression^7–12^, suggesting that ctDNA contains accurate and quantifiable information for longitudinally tracking tumor progression.

Specifically, many assays quantify the abundance of somatic mutations via the variant allele fraction (VAF); however, tumor-naive assays that rely on quantifying somatic mutations face technical limitations for precise therapy response monitoring. First, a significant fraction of advanced cancer patients (up to 20%) may not have any somatic variants detected in plasma^12–14^, rendering the assay unusable for therapy response monitoring. If variants are detected, the number of present variants can be limited (a mean between 3 and 4)^10, 13^, and the VAF is often small (<0.5%)^13, 15^, reflecting a low abundance of total variant molecules. This results in a high coefficient of variation (CV) due to Poisson sampling alone. For example, a single variant at 0.5% VAF in a sample with 2000 genomic equivalents would contain an average of 10 variant molecules, resulting in a minimum CV of 32% even with 100% assay quantification accuracy. A high CV means that each individual measurement has a high amount of noise and variability, making longitudinal measurements difficult to interpret, and potentially results in spurious apparent changes in ctDNA abundance simply due to random sampling. To better ensure the detection of a sufficient number of somatic variants and variant molecules, tumor-informed assays have been developed, in which a personalized set of somatic mutations are identified from a tumor biopsy and then assayed in a subsequent liquid biopsy^16–19^. However, when implementing this two-step approach for assessment of therapy response, the procedure of obtaining a biopsy is invasive, and sometimes impossible, while simultaneously prolonging turnaround time. Moreover, tumor-informed assays track the abundance of a limited number of somatic mutations, whose abundance may not accurately reflect the tumor composition especially when the tumor clonal heterogeneity changes under pressure from a therapy, as is often true in the case of late-stage cancers^20^.

Given that precision is often limited by the Poisson sampling noise associated with assaying somatic mutations, methylation of ctDNA has been explored as an alternative biomarker to VAF measurements of ctDNA. Generally, methylation has been shown to be a strong, consistent, and genomically widespread biomarker for cancer that can also be detected in ctDNA^21, 22^ . While methylation signals are more abundant compared to somatic mutations, quantifying the amount of methylation accurately and precisely is a challenge. Methylation-specific qPCR has been shown to be correlated with clinical outcomes^23–25;^ however, because of its exponential nature, qPCR has high assay noise when calculating the absolute number of molecules, typically in the tens of percent^26^. Sampling hundreds of, instead of one or two, genomic locations for methylation would significantly improve the assay performance, but interrogating multiple loci using qPCR is technically infeasible without obtaining additional cfDNA, i.e. additional tubes of blood for each locus, from the patient. Current DNA methylation sequencing methods also face challenges with precise quantification^27^. Therefore, a new technical approach is needed to precisely quantify ctDNA methylation while taking advantage of its abundance in the genome.

Herein, we describe Northstar Response, a novel assay that uses molecule counting to accurately and precisely quantify methylated ctDNA for tumor-naive cancer therapy response monitoring. We built a multiplex assay that targets more than 500 locations in the genome that are hypermethylated in cancer compared to normal tissue and leveraged quantitative counting templates (QCTs)^28^ to count the number of methylated molecules at each of these locations.

We analytically validated this assay, accurately distinguishing 0.25% absolute changes in tumor fraction (AUC > 0.94) and demonstrating high accuracy across 12 tumor tissue types. We show that this assay is not only correlated with but also up to two times more precise than comparable VAF-based therapy response monitoring methods, achieving CVs <10% in 1% tumor fraction samples. Finally, we show high concordance between clinical outcomes from cancer patients and Northstar Response methylation measurements.

## Results

### Design of Northstar Response

We designed a pan-cancer, amplicon-based, multiplex PCR assay that utilizes QCTs in conjunction with next-generation sequencing to count the number of methylated molecules at more than 500 genomic locations known to be hypermethylated in cancer tissue compared to normal tissue (Figure 1A). To identify these hypermethylated locations, we queried The Cancer Genome Atlas (TCGA) for subjects with human methylation data for both tumor and normal tissue of the same tissue type^29^. Tumor hypermethylation was calculated at each CpG site by subtracting normal tissue beta values from tumor beta values, where beta represents the ratio of the methylated array intensity to the total array intensity. Tumor hypermethylation was averaged across all CpG sites in the same CpG island to remove spuriously methylated CpG sites and ranked in order of highest hypermethylation across 12 different tumor tissue types (Figure 1D). Additionally, CpG islands with an average white blood cell beta < 0.2, measured from patients of similar age to cancer patients (mean = 63.9 years, sd = 13.3 years)^30^, were filtered out to minimize background signal from buffy coat contributions to the cfDNA. In addition to selecting targets that are hypermethylated in tumors compared to normal tissue, CpG sites that are highly methylated in all tissues (e.g., buffy coat, tumor, and normal tissue) were selected to estimate the total amount of input DNA for downstream normalization. These normalization loci were selected using the same methylation data sources: CpG sites were ranked in order of highest white blood cell methylation signal and filtered for average beta > 0.9 across CpG islands for white blood cell, tumor tissue, and normal tissue for six tumor tissue types.

**Figure 1.**
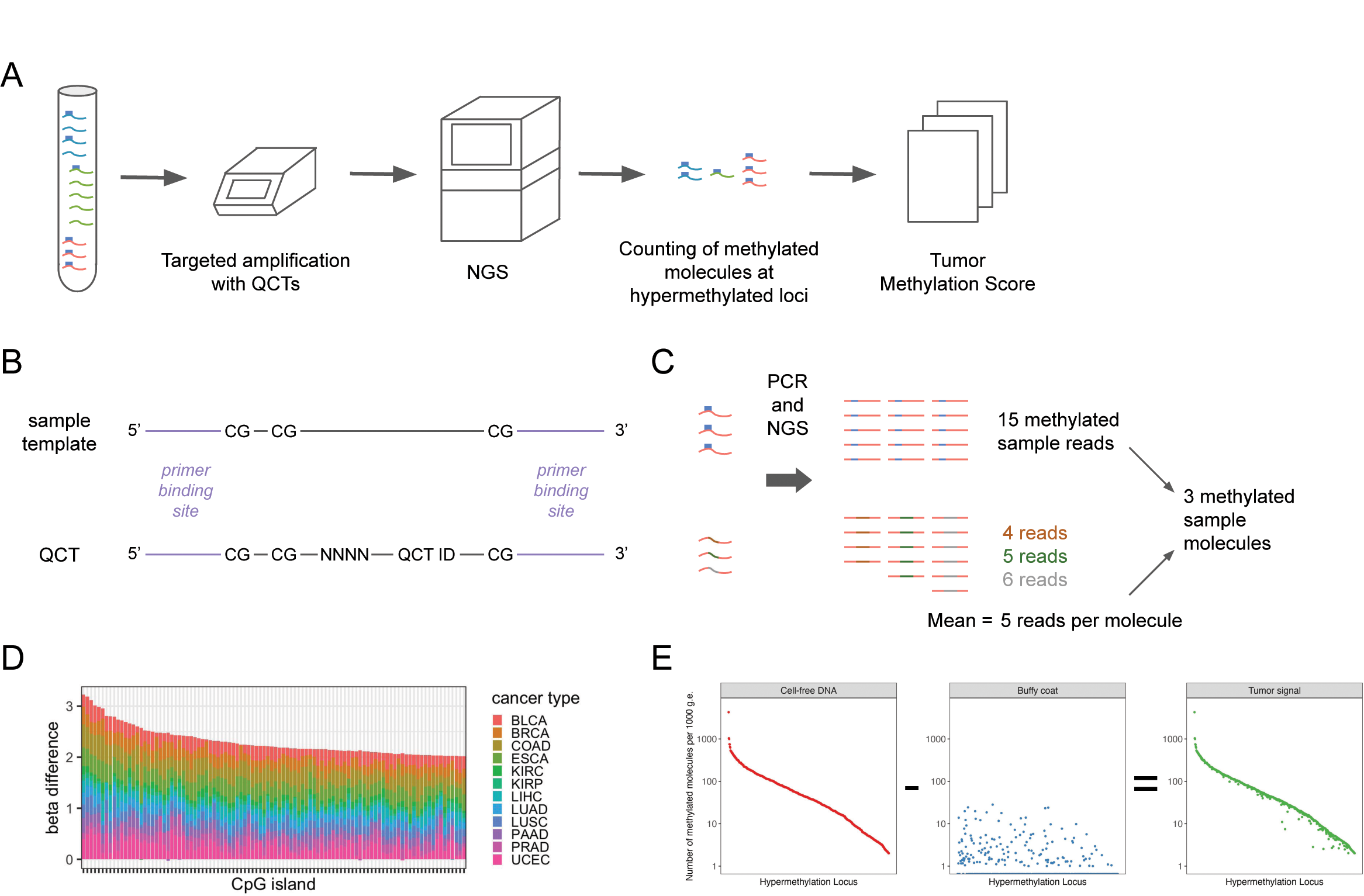
Design and analysis for Northstar Response. (**A**) Workflow overview for Northstar Response. cfDNA (cell-free DNA) molecules are hypermethylated (blue rectangles) in ctDNA (circulating tumor DNA) compared to normal tissue at multiple genomic locations (red, green, and blue wavy lines). These cfDNA molecules at targeted hypermethylated loci are co-amplified with Quantitative Counting Templates (QCTs) and sequenced using next-generation sequencing (NGS). This data is analyzed to calculate the number of methylated molecules at these targeted loci and aggregated across all loci to calculate the Tumor Methylation Score. (**B**) QCTs are designed for each targeted genomic location such that they have identical primer binding site sequences but have a number of random bases that determine an embedded molecular identifier (EMI) and a QCT identification sequence to distinguish QCTs from sample molecules. (**C**) The number of reads per EMI is averaged across all EMIs for that genomic location. Because QCTs are added at an abundance ensuring one molecule per EMI, and that QCTs amplify at the same rate as sample molecules at the targeted genomic location, the number of reads per EMI is the number of reads per molecule at that genomic location. The number of methylated sample reads can then be divided by the number of reads per molecule to calculate the number of methylated sample molecules at the start of PCR. (**D**) The top 100 CpG islands ranked by total hypermethylation across 12 cancer types according to TCGA data. Beta is the fraction of DNA that is methylated, and the beta difference was calculated by subtracting the beta measured in normal tissue from the beta measured in tumor tissue. Included cancer types were bladder urothelial carcinoma (BLCA), breast invasive carcinoma (BRCA), colon adenocarcinoma (COAD), esophageal carcinoma (ESCA), kidney renal clear cell carcinoma (KIRC), kidney renal papillary cell carcinoma (KIRP), liver hepatocellular carcinoma (LIHC), lung adenocarcinoma (LUAD), lung squamous cell carcinoma (LUSC), pancreatic adenocarcinoma (PAAD), prostate adenocarcinoma (PRAD), and uterine corpus endometrial carcinoma (UCEC) (**E**) The numbers of methylated molecules measured in paired cfDNA and buffy coat are first normalized to the estimated input genomic equivalents (g.e.), then background methylation detected in buffy coat is subtracted from the methylation measured in plasma on a per-locus basis to extract tumor-associated signal.

In addition to designing amplicons and their associated primers for selected CpG sites, corresponding QCTs were designed for each target amplicon to estimate the input number of methylated molecules at each locus through next-generation sequencing data analysis (Figure 1B, 1C). To account for varying input amounts, the number of methylated molecules is normalized to the estimated input amount of DNA as measured by the average number of methylated molecules across highly methylated normalization loci. As a significant portion of cfDNA signal is of white blood cell origin^31^, we can reduce background methylation by subtracting the number of methylated molecules measured in the buffy coat from that measured in the paired cfDNA on a per-locus basis (Figure 1E, Supplementary Figure 8). This background subtraction step lowers the noise floor of the assay and likely also minimizes spurious methylation signals resulting from effects that are non-specific to the cancer, such as the side effects of systemic therapy. The result is a per-locus number of tumor molecules which is summed across loci to calculate the Tumor Methylation Score for the sample.

### Quantification of tumor DNA with Northstar Response

To assess the analytical performance of the Northstar Response assay, we tested replicates of contrived samples made from sheared tumor DNA spiked into paired sheared buffy coat DNA at 0%, 0.25%, 0.5%, 1%, and 2% tumor fraction by mass. By assaying contrived technical replicates, we were able to perform repeated measurements of Tumor Methylation Score on samples with known amounts of tumor DNA. This experiment was repeated using two distinct tumors, a stage IV lung adenocarcinoma and a stage IV breast adenocarcinoma. For both tumors, the Tumor Methylation Score linearly increased with contrived tumor fraction, accurately quantifying the amount of methylated tumor DNA in contrived samples (breast *R*^2^ = 0.998, lung *R*^2^ = 0.996) (Figure 2A). Notably, this relationship is directly proportional, with a doubling in contrived tumor fraction closely corresponding to a doubling in Tumor Methylation Score. When the background subtraction used to calculate Tumor Methylation Score is not performed, this proportionality is lost due to background signal interference (Supplementary Figure 8A).

**Figure 2.**
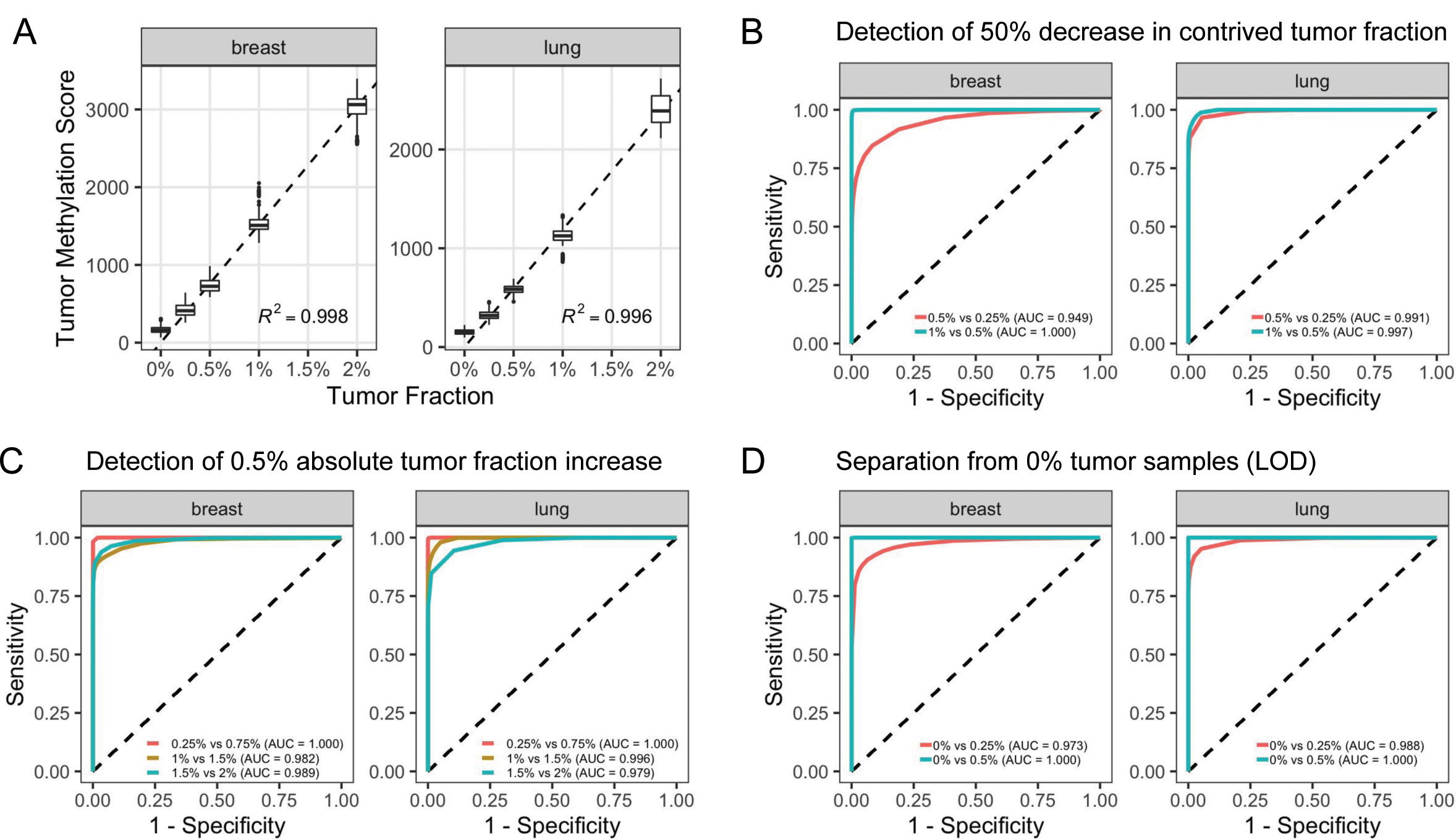
Northstar Response accurately and precisely quantifies methylated ctDNA. (**A**) Two tumors were used to generate technical replicates of contrived samples at different tumor fractions by mass, assuming that the tumor was 100% cancerous tissue (total input mass = 30 ng). The replicates were processed independently through the Northstar Response assay, and then the Tumor Methylation Score was calculated for every possible combination of contrived sample and buffy coat. The boxplots show the median and interquartile range in Tumor Methylation Score for each contrived tumor fraction. The dashed lines indicate a linear fit through the origin and the mean Tumor Methylation Score for each contrived tumor fraction. (**B**) Receiver operating characteristic (ROC) curves for the sensitivity of calling changes in Tumor Methylation Score after a halving of contrived tumor fraction. Specificity was measured for comparisons of the larger tumor fraction against itself. (**C**) ROC curves for calling changes in Tumor Methylation Score after an increase of 0.5% contrived tumor fraction. Data for 0.75% and 1.5% tumor fraction were obtained by appropriately scaling down the signal from 1% and 2% replicates, respectively. (**D**) ROC curves for calling changes from Tumor Methylation Score of 0% tumor fraction to Tumor Methylation Score from contrived 0.25% and 0.5% tumor fraction replicates and specificity of calling no change for 0% vs 0% tumor fraction comparisons. AUC, area under the curve. LOD, limit of detection.

Additionally, the CV of the Tumor Methylation Score was below 20% for all contrived tumor fractions tested, decreasing to about 6% at 2% tumor fraction (Figure 3). These results demonstrate that Northstar Response quantifies methylated tumor DNA with high accuracy and precision.

**Figure 3.**
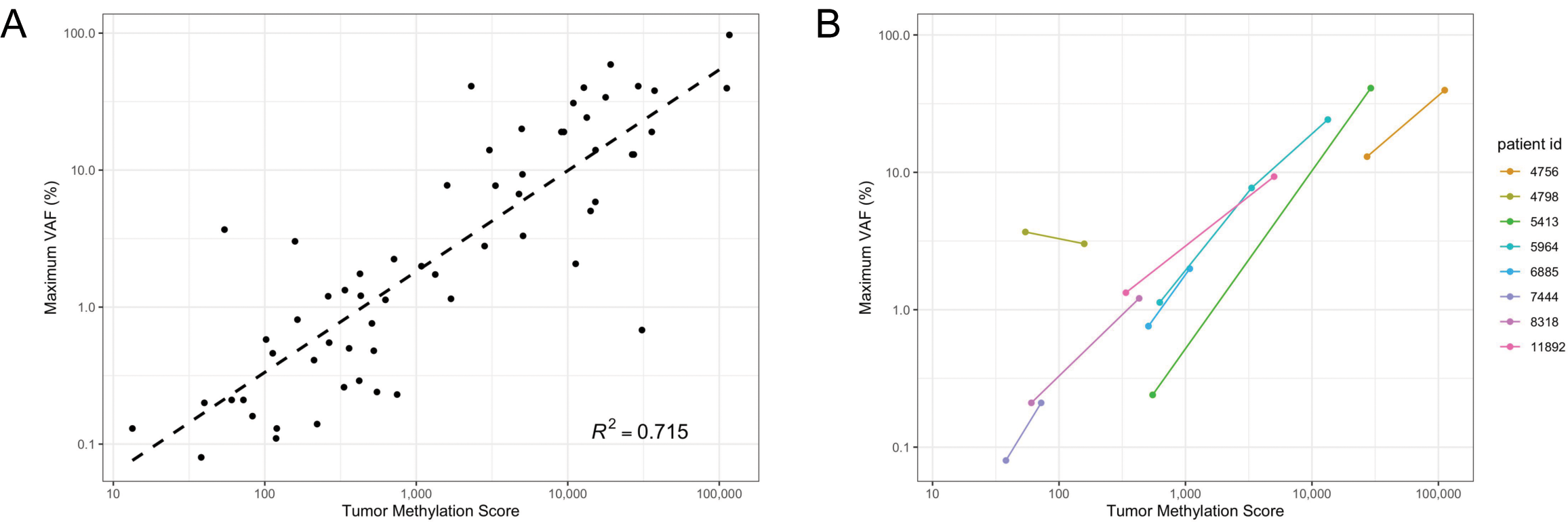
Correlation of Tumor Methylation Score with VAF in clinical samples from metastatic lung cancer patients. (**A**) Correlation of maximum VAF among clinically significant variants as measured by a treatment selection assay with Tumor Methylation Score in clinical samples (N = 60 samples). Mutations of clonal hematopoiesis of indeterminate potential (CHIP) were excluded based on results from testing the matched buffy coat sample. A linear fit and corresponding *R*^2^ was calculated using the logarithm of maximum VAF and the logarithm of Tumor Methylation Score. (**B**) Correlation of Tumor Methylation Score with maximum VAF within patients for therapy response monitoring (N = 8 patients). CHIP mutations were similarly excluded.

### Classification of changes in tumor fraction using Tumor Methylation Score

Because Northstar Response is intended to monitor changes in methylated ctDNA abundance over time, we also evaluated the assay’s ability to distinguish between different contrived tumor fractions. Based on the measured Tumor Methylation Scores of a given pair of samples, the assay made calls to determine whether there was a change in contrived tumor fraction from the first sample to the second sample. We generated receiver operating characteristic (ROC) curves for different comparisons by calculating the sensitivity and specificity of calls starting at a given contrived tumor fraction over a range of calling thresholds. Overall, the assay was able to call changes in Tumor Methylation Score with high sensitivity and specificity (Figure 2B-D).

Previous studies have shown that molecular response to immunotherapy, defined as a >50% decrease in mean VAF, is associated with favorable clinical outcomes^7, 10, 12^. Starting at a contrived tumor fraction of 1%, Northstar Response was able to distinguish whether there was a decrease to 0.5% tumor fraction or no change in tumor fraction with accuracy approaching 100% (Figure 2B). For a 50% decrease from 0.5% to 0.25% contrived tumor fraction, the area under the ROC curve (AUC) was 0.949 for the breast tumor and 0.991 for the lung tumor. The assay performed similarly for a doubling in contrived tumor fraction, making correct calls for tumor fraction increases as small as from 0.25% to 0.5% tumor fraction (breast AUC = 0.918, lung AUC = 0.981) (Supplementary Figure 1B).

While large reductions in measured ctDNA are most strongly associated with improved survival, capturing smaller changes in methylated ctDNA levels may provide additional clinical utility by enabling earlier detection of progression. The assay made accurate calls for step increases of 0.5% contrived tumor fraction, with AUC > 0.97 for all comparisons tested (Figure 2C). Although distinguishing between samples with smaller step increases of 0.25% tumor fraction was slightly more challenging, particularly as the relative change between the samples becomes small, the assay could still distinguish between 1.75% tumor and 2% contrived tumor fraction (breast AUC = 0.818, lung AUC = 0.727) (Supplementary Figure 1C).

We also investigated the assay’s limit of detection, defined as the lowest tumor fraction that could be distinguished from 0% tumor samples with 95% sensitivity. The assay differentiated well between 0% and 0.25% tumor samples for both tissue types (breast AUC = 0.973, lung AUC = 0.988) (Figure 2D). While maintaining at least 90% specificity, the assay achieved 92.4% sensitivity for the breast tumor and 95.2% sensitivity for the lung tumor. Variability in the performance between the two samples is likely due to differences in the level of background signal found in the paired buffy coat (Supplementary Figure 1A). For the comparison between 0% and 0.5% tumor samples, the assay made calls with 100% sensitivity for both tumor types (AUC = 1.000). These results indicate that the assay’s limit of detection is between 0.25% and 0.5%.

### Correlation of Tumor Methylation Score with VAF in clinical samples

While the above results demonstrate that Northstar Response accurately quantifies the amount of ctDNA in contrived samples, we also wanted to show that Tumor Methylation Score measurements reflect the amount of ctDNA in clinical plasma samples. To obtain orthogonal measurements of ctDNA abundance, paired plasma samples originating from a single blood draw were processed using both Northstar Response and a treatment selection assay that detects somatic mutations. Paired buffy coat samples from each patient were also processed with both Northstar Response and the VAF-based assay, enabling both subtraction of background methylation signal and removal of somatic mutations arising due to clonal hematopoiesis of indeterminate potential (CHIP), respectively. Within this patient cohort, there is a positive correlation between maximum VAF and Tumor Methylation Score (*R*^2^ = 0.715, log-log regression) (Figure 3A). When mutations attributed to CHIP are included in the analysis, the amount of unexplained variance in the data increases (*R*^2^ = 0.598, log-log regression) (Supplementary Figure 2A), indicating that CHIP is another source of noise and error for quantifying ctDNA level in VAF-based assays that do not sequence the buffy coat. We also investigated the concordance between Northstar Response and the VAF-based assay in a therapy response monitoring context. A total of ten patients had samples available from multiple time points; however, two of these patients had only CHIP-derived somatic mutations detected at one or more time points, making it impossible to accurately track ctDNA dynamics with the VAF-based assay. We observed consistent trends in maximum VAF and Tumor Methylation Score measurements for 7 out of 8 patients with somatic mutations detected (Figure 3B, Supplementary Figure 2B). These results indicate that Northstar Response detects ctDNA-specific methylation that correlates with VAF-based measurements of ctDNA abundance.

### Comparing Northstar Response precision with a VAF-based therapy response monitoring approach

To evaluate how the precision of the Northstar Response assay compares to a VAF-based monitoring approach, we compared the CV of Tumor Methylation Score to the CV of VAF of somatic mutations measured using a hybrid capture-based treatment selection assay. A contrived sample with DNA from multiple tumors was diluted into healthy background DNA at 10% and 20% tumor material by mass, with 20 replicates of each condition, to generate samples with many mutations at a wide range of VAFs. Average VAF was calculated for each mutation across the 20 replicates and was used to filter and group the mutations into 0.25%, 0.5%, 1%, and 2% tumor fraction bins. More than half of liquid biopsy samples have four mutations or fewer^13^; therefore, we generated scenarios where either one, two, three, or four mutations were detected in a patient cfDNA sample and the VAFs for those variants were then averaged for therapy response monitoring. We selected all possible combinations of one, two, three, and four mutations from each tumor fraction bin, averaged the VAFs across mutations, and calculated the coefficient of variation of these average VAFs. To validate these empirical results, a Poisson sampling-based simulation was performed assuming mutations at 0.25%, 0.5%, 1%, and 2% VAF to calculate the theoretical CV. These empirical and theoretical results were compared to the CV of Tumor Methylation Score in replicates of breast and lung tumor-derived contrived samples at matching contrived tumor fractions of 0.25%, 0.5%, 1%, and 2%.

The CV of Tumor Methylation Score for both the breast and lung tumor contrived samples was lower than the median CV and simulated CV of mean VAF at all tested VAF tumor fraction bins (Figure 4). In fact, Northstar Response achieved CVs twice as low as that measured using a VAF-based approach, particularly when few mutations are measured using the VAF-based approach. Even if we assume no additional noise beyond Poisson sampling noise (e.g., no PCR or sequencing error, perfect molecular capture, and no clonal heterogeneity), a VAF-based approach would need to detect at least 10 somatic mutations to match the precision of Northstar Response for a 2% tumor fraction sample (Supplementary Figure 3). Of note: the mean number of somatic mutations that can be tracked in advanced cancer patients using a commonly used tumor-naive VAF-based assay is between 3 and 4 mutations^10, 13^.

**Figure 4.**
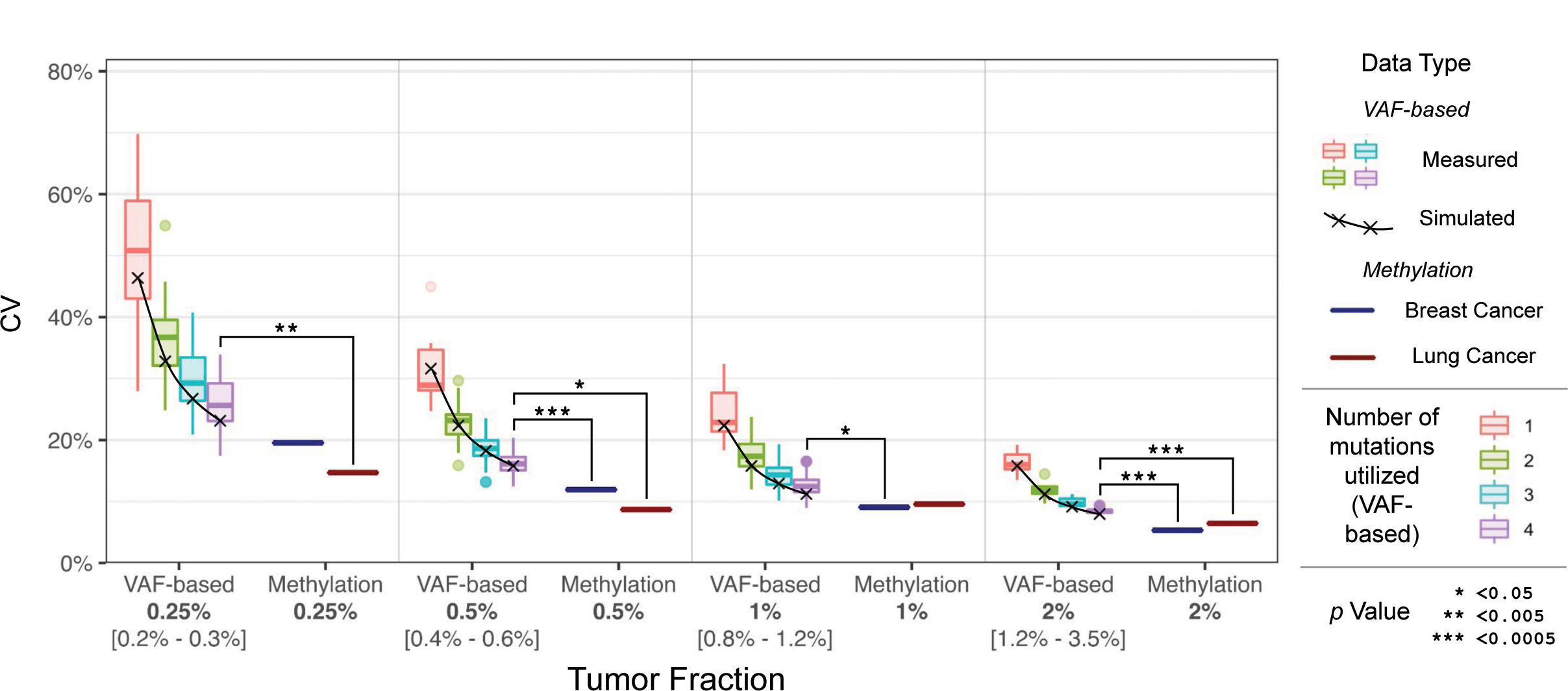
Coefficients of variance for VAF-based and methylation-based therapy response monitoring. VAF-based CVs were calculated through empirical measurements (boxplots) and simulation (black X’s). CV of Tumor Methylation Scores were calculated at matching contrived tumor fractions (dark blue and red bars). *p* values for each tumor fraction were calculated comparing the Methylation Score CV for each cancer type to the distribution of average VAF CVs for 4 mutations utilized using a 2-sided T test (black asterisks).

### Analytical validation of Northstar Response in several different tissues of tumor origin

While Northstar Response accurately and precisely quantified DNA methylation in two tumors, we also tested its performance across a more expansive set of tumors. Using 54 solid tumors originating from 12 different tissues, we tested contrived samples that were made by combining sheared tumor DNA and matched sheared buffy coat DNA at 0%, 1%, and 2% tumor fraction. To account for the variable fraction of cancerous tissue in each specimen, we calculated a tumor purity score for each tumor and scaled tumor DNA inputs accordingly when preparing contrived samples (Supplementary Figure 4).

Tumor Methylation Scores at 1% and 2% tumor fraction were calculated for each tumor by subtracting background methylation detected in 0% tumor samples. The Tumor Methylation Scores for all 2% tumor fraction samples were 1.65x - 2.35x greater than those of the 1% tumor fraction samples (Figure 5A), except for one lung neuroendocrine tumor for which we measured little difference in signal likely due to low tumor purity (Supplementary Table 2). Quantifying the fold change in methylation without accounting for background signal detected in buffy coat resulted in less accurate and more variable measurements (Supplementary Figure 8B, 8C), which highlights the utility of subtracting background methylation when calculating Tumor Methylation Scores. The 2% tumor samples were then called as having increased, unchanged, or decreased Tumor Methylation Scores relative to 1% tumor samples. We correctly made increase calls for 53/54 sample pairs, which corresponds to 98.1% [95% confidence interval: 90.11%, 99.95%] sensitivity across tissue types. While the assay is able to accurately call relative increases from 1% to 2% contrived tumor fraction within individual patients, we observed variability when comparing Tumor Methylation Score between patients and across tissue types measured at a given contrived tumor fraction (Supplementary Figure 5A).

**Figure 5.**
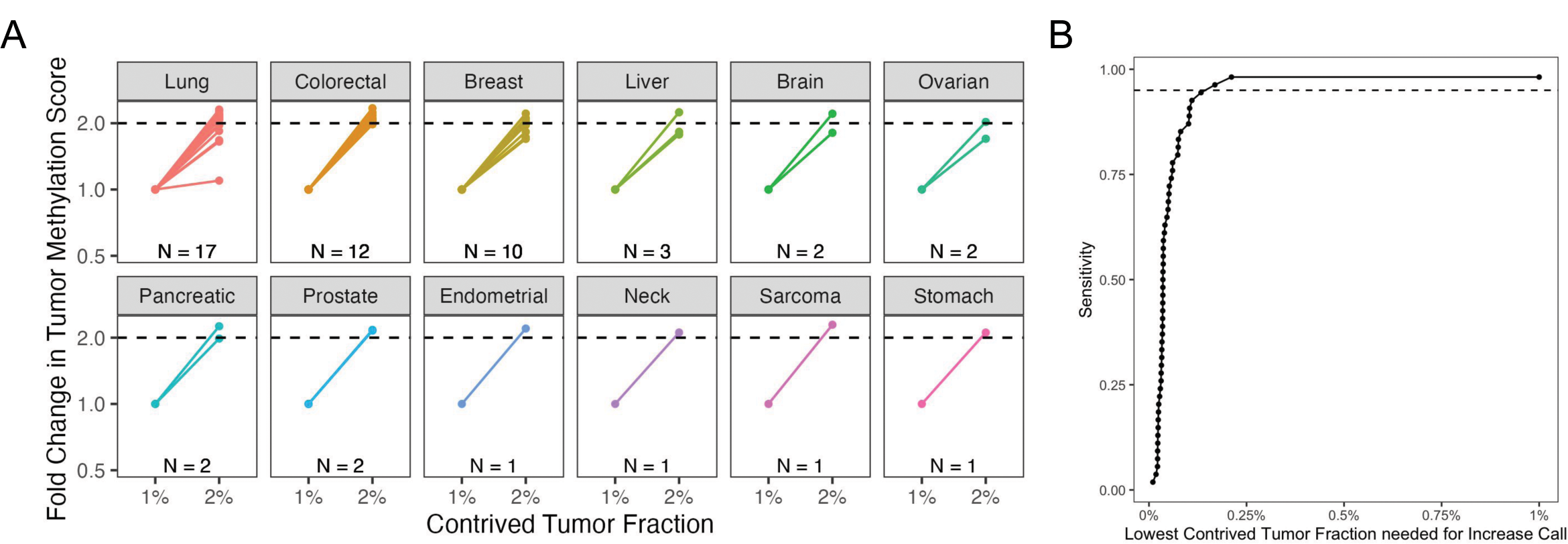
(**A**) Tumor Methylation Scores were significantly increased at 2% tumor fraction relative to 1% tumor fraction for 53/54 different tumor specimens across 12 different tissues of origin. (**B**) The lowest contrived tumor fractions at which increase calls could have been made were estimated by linearly scaling down Tumor Methylation Scores detected at 1% and 2% tumor fraction i*n silico*, maintaining a doubling in contrived tumor fraction. At contrived tumor fractions as low as 0.17%, increase calls could be made for 52/54 tumor specimens, which corresponds to 96.3% sensitivity.

Since the assay detected increases in contrived tumor fraction from 1% to 2% with high confidence for the 53 tumors called correctly, we also simulated assay sensitivity at lower tumor fractions. To do this, we linearly scaled down Tumor Methylation Scores detected at 1% and 2% tumor fraction *in silico*, maintaining a doubling in contrived tumor fraction. For each tumor, we then identified the lowest contrived tumor fraction at which we would have still made an increase call. We found that increase calls would still be made with 98.1% sensitivity (53/54 correct calls) when contrived tumor fractions increase from 0.21% to 0.42% and with 96.3% sensitivity (52/54 correct calls) when contrived tumor fractions increase from 0.17% to 0.34% (Figure 5B).

### Clinical results from Northstar Response

To investigate whether the Northstar Response assay could measure clinically meaningful signals, we tested cancer patient samples collected before and after treatment. Tumor Methylation Scores were determined at each sample collection time point, and calls were made by comparing the Tumor Methylation Score with the immediately preceding measurement. These longitudinal results were compared with clinic-reported clinical outcomes that were determined from imaging scans. A total of seven patients had at least three sample collection time points with at least one corresponding clinical outcome within 100 days of any sample collection time point. These patients all had advanced stage cancer, and their cancers spanned three different tissues of origin: lung, pancreas, and colorectal cancers. Additional demographic information can be found in Supplementary Table 1.

We observed changes in Tumor Methylation Scores that reflected the dynamics of cancer treatment (Figure 6). All seven patients had an initial decrease in Tumor Methylation Score after starting treatment (Figure 6H). Some of these decreases were large, with one patient’s Tumor Methylation Score decreasing by almost 80% within 7 days (Figure 6F) and several patients’ Tumor Methylation Scores decreasing by more than 90% compared to pre-treatment levels (Figure 6A-E, 6G). We also observed large increases in Tumor Methylation Score later in the course of treatment, with some patients increasing between 4-fold and 160-fold (Figure 6A-E). Changes as small as 2-fold were also observed (Figure 6A, 6F), and sometimes, the Tumor Methylation Score remained below the noise floor in consecutive time points (Figure 6E, 6G).

**Figure 6.**
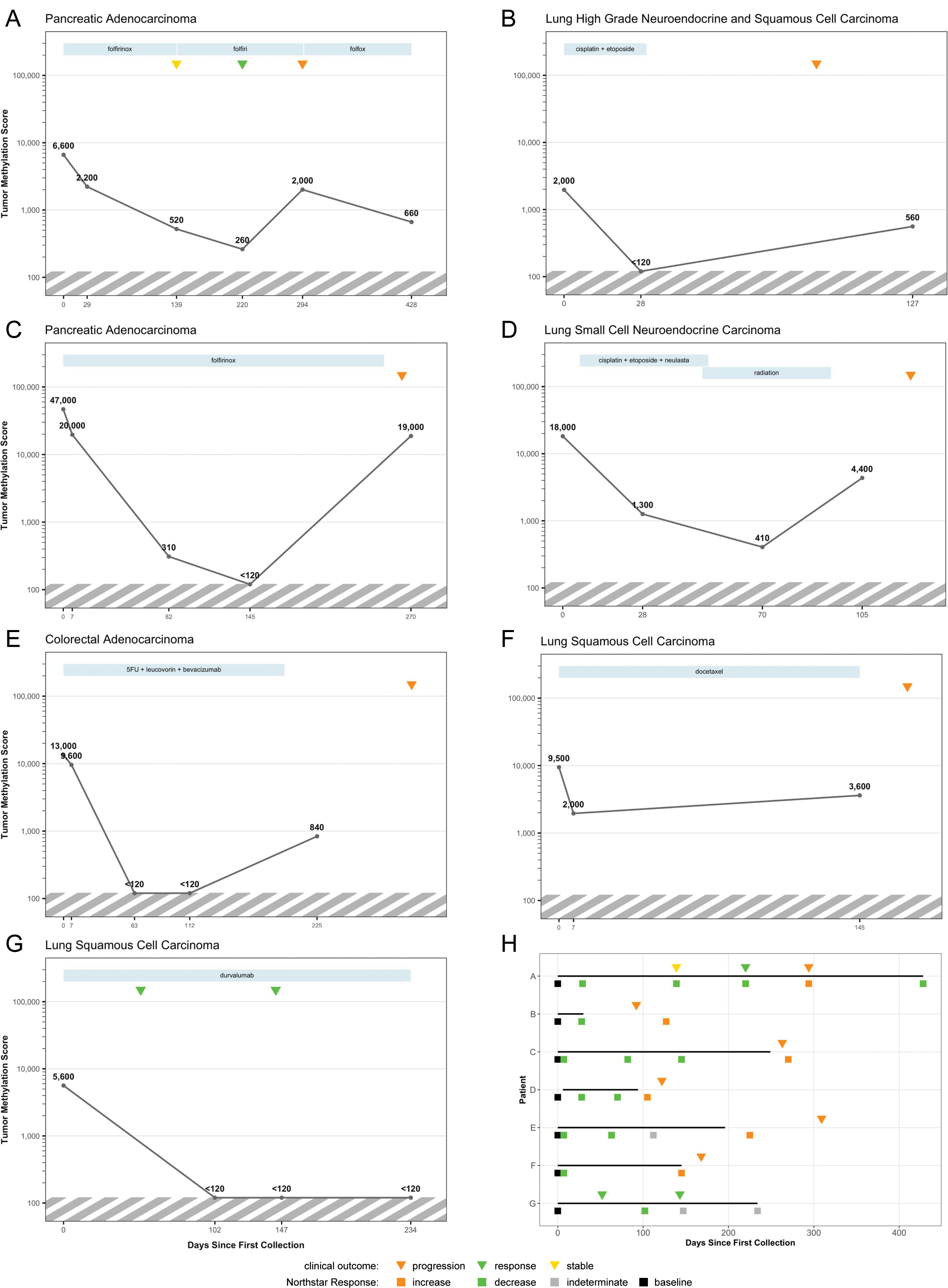
Longitudinal clinical results from Northstar Response. (**A-G**) Tumor Methylation Score measurements from seven cancer patients, annotated with clinical outcomes indicated by triangles (orange = progression, green = response, yellow = stable) as reported by the clinic. Blue rectangles plotted at the top indicate the duration of each treatment regimen. The striped region on each plot indicates the noise floor; any Tumor Methylation Scores below 120 are reported as <120. (**H**) Swimmer plot summarizing Tumor Methylation Score and clinical outcomes for all seven cancer patients. Squares indicate calls made by Northstar Response based on changes in Tumor Methylation Score compared to the previous sample point (orange = increase, green = decrease, gray = indeterminate, black = baseline measurement). Segments indicate the duration of treatment.

Tumor Methylation Scores showed high concordance with clinical outcomes. Of the six patients that displayed an increase in Tumor Methylation Score, all six had progression noted as a clinical outcome shortly before or shortly after the sample was collected (Figure 6H). In three of those six patients, an increase in Tumor Methylation Score was measured before clinical progression was noted (Figure 6D, 6E, 6F). In addition, when a partial response clinical outcome was noted, the Tumor Methylation Score had decreased compared to the pre-treatment time point (Figure 6A, Figure 6G). In one of these patients, Tumor Methylation Scores had decreased by over 10-fold 81 days prior to a reported partial response clinical outcome (Figure 6A). Taken together, these results suggest that Northstar Response may be a useful tool for therapy response monitoring in the clinical setting.

## Discussion

Therapy response monitoring through liquid biopsy has been increasingly shown to be clinically useful for late-stage cancer patients. However, current liquid biopsy approaches for therapy response monitoring face several technical challenges that limit both the accuracy and precision of those measurements. In this study, we described Northstar Response, a novel, tumor-naive, methylation-based assay, that overcomes the aforementioned technical challenges.

The assay is highly accurate and precise and has high sensitivity in multiple tumors from different tissues of origin. At 2% contrived tumor fraction, the CV of the measured Tumor Methylation Score is less than 6.5%. This level of precision enables the detection of 20% changes in Tumor Methylation Score with 99.7% confidence (an interval of 3 standard deviations), meaning that the assay could call an increase in ctDNA burden from 2% to 2.4% contrived tumor fraction or a decrease from 2% to 1.6% tumor fraction. While the clinical significance of small changes in ctDNA burden remains to be elucidated within the field of liquid biopsy, the ability to detect these changes may provide earlier insight into a patient’s response to therapy compared to traditional imaging methods. Further studies of Northstar Response will be required to establish the appropriate thresholds for molecular progression and response based on changes in Tumor Methylation Score.

To further characterize Northstar Response’s ability to quantify ctDNA in clinical samples, we processed patient samples using both Northstar Response and an orthogonal VAF-based treatment selection assay. We found that the Tumor Methylation Score measured by Northstar Response was correlated with maximum VAF, and both assays measured concordant trends in ctDNA dynamics across multiple time points from the same patient. These results show that in addition to high analytical sensitivity and specificity on contrived samples, Northstar Response accurately quantifies ctDNA in clinical samples for therapy response monitoring. In this set of patient samples, we also observed that the inclusion of CHIP-derived variants weakened the correlation between maximum VAF and Tumor Methylation Score. While the false positive ctDNA signal from CHIP-associated mutations can be filtered out by processing the paired buffy coat sample, commercially available VAF-based assays often omit this step. Furthermore, if CHIP-derived mutations are excluded from analysis, the fraction of patients with no somatic mutations detected may increase. Northstar Response’s methylation-based approach overcomes these challenges faced by VAF-based liquid biopsies, both of which can significantly interfere with therapy response monitoring.

VAF-based assays are currently used for therapy response monitoring by tracking the relative abundance of somatic mutations over time. However, the precision of tumor-naive VAF-based assays is often limited by Poisson counting noise due to the small number of detectable variants, making these assays poorly suited for therapy response monitoring. This lack of precision could misclassify patients as molecular responders / non-responders, such as when predicting clinical outcomes for immunotherapy. We found that Northstar Response consistently achieved lower CV and thus greater precision compared to using a VAF-based measurement for therapy response monitoring. One reason for this improved performance is that the abundance of informative methylated loci is much greater than the typical number of somatic variants detected in a tumor-naive assay (Supplementary Figure 7), which significantly reduces the Poisson sampling noise of Northstar Response. The improved precision allows for the detection of smaller changes in the abundance of ctDNA, which could translate to measuring signs of tumor progression, or response, earlier.

Northstar Response maintained accuracy and sensitivity across a diversity of tumors and patients, suggesting that methylation-based therapy response monitoring has the potential to benefit a broad intended use population. Northstar Response correctly detected increases from 1% to 2% tumor fraction with 98.1% sensitivity for 54 solid tumors from 12 distinct tissue types. Despite the heterogeneity often observed in cancer biology, this implies that sampling a panel of hypermethylated loci enables pan-cancer and tumor-naive tracking of tumor progression. Especially for tumors present in tissues that are difficult to image such as bone and the peritoneum, Northstar Response may be a useful alternative for evaluating therapy response.

In addition to high performance for distinguishing 1% from 2% tumor fraction, simulating lower contrived tumor fractions revealed that Northstar Response is able to detect minute changes in tumor fraction for a variety of tissue types and tumor stages. In particular, Northstar Response maintained 98.1% sensitivity for detecting increases in contrived tumor fraction from 0.21% to 0.42%, demonstrating the ability to resolve small changes in the amount of tumor DNA for many tissue types. In the future, additional tumor samples from untested tissues of origin should be tested to further validate Northstar Response as an effective therapy response monitoring test for all solid tumors.

To further assess the clinical validity of Northstar Response, we tested Northstar Response in longitudinally collected samples from cancer patients, and we found that Tumor Methylation Scores correlated strongly with clinical outcomes. Notably, the assay detected a significant increase in Tumor Methylation Score in advance of progression in three patients, suggesting the possibility of detecting molecular progression before radiologically documented progression. The timing of both the blood sample collection and clinical outcome collection were variable in this study; a clinical study with regimented sample and imaging data collection times may better determine how early and how consistently the assay could detect progression before imaging. In practice, the clinical utility of the assay is best demonstrated in measuring molecular progression whereby the oncologist may consider accelerating the timing of the next imaging scan and adjust treatment regimens accordingly. Future interventional clinical utility studies will be needed to assess whether making such changes based on a liquid biopsy result will ultimately result in improved clinical outcomes.

Northstar Response also detected decreases in Tumor Methylation Score corresponding with partial response clinical outcomes, and in one case, detected a decrease in advance of an imaging clinical outcome. The potential of Northstar Response to detect molecular response before imaging, in addition to giving patients peace of mind that their therapy is working, could also help clinicians predict clinical outcomes further in advance. While previous VAF-based therapy response monitoring studies have shown that patients receiving immunotherapy^7, 10– 12, 19, 32, 33^ or targeted therapy^8, 34^ that achieve molecular response have improved outcomes, the improved precision of Northstar Response may enable the accurate detection of smaller yet still meaningful molecular response. Clinical studies will be needed to test this hypothesis.

While ctDNA-based assays have advantages compared to imaging for therapy response monitoring, such as improved accessibility to patients and the ability to detect smaller total amounts of tumor, we imagine that the information from therapy response monitoring assays would complement imaging scans performed as part of the standard of care by adding an independent and incremental measurement of the cancer. Critically, imaging can identify the presence and spatial location of metastases; it remains to be seen whether biomarkers in the ctDNA like methylation could detect such metastases and distinguish them from the primary tumor. Overall, Northstar Response is a technical advance that may improve therapy response monitoring and enhance cancer patient care.

## Methods

### Target selection

To choose targets that are hypermethylated in tumors, we queried The Cancer Genome Atlas (TCGA) for subjects with methylation data from matched tumor and normal tissue of the following cancer types: colon adenocarcinoma (COAD), liver hepatocellular carcinoma (LIHC), lung adenocarcinoma (LUAD), lung squamous cell carcinoma (LUSC), breast invasive carcinoma (BRCA), pancreatic adenocarcinoma (PAAD), bladder urothelial carcinoma (BLCA), esophageal carcinoma (ESCA), kidney renal clear cell carcinoma (KIRC), kidney renal papillary cell carcinoma (KIRP), prostate adenocarcinoma (PRAD), and uterine corpus endometrial carcinoma (UCEC). To choose targets for normalization loci that are highly methylated in buffy coat, tumor, and normal tissue, methylation data was obtained and analyzed from six tumor tissue types: lung adenocarcinoma, lung squamous cell carcinoma, colon adenocarcinoma, breast invasive carcinoma, pancreatic adenocarcinoma, and liver hepatocellular carcinoma.

### Oligo design and preparation

Primer3 (https://primer3.org/) was used to design primer pairs targeting the genomic locations of chosen CpG locations and using a reference genome that was bisulfite converted *in silico*. This process was performed assuming full methylation of the reference hg19 human genome, converting all Cs to Ts except for CpGs. The ideal annealing temperature was set to 60C. Primers were ordered from Eurofins Genomics (Louisville, KY).

Primer mixes were created by pooling all primer pairs and iteratively removing and/or rebalancing the concentrations of each primer pair to optimize for balanced read depth across target amplicons. Two versions of the assay were created; a v0 assay with 590 total amplicons and a v1.1 assay with 551 total amplicons. The v0 assay was used for testing clinical samples, and the v1.1 assay was used for all other presented results.

Single stranded Quantitative Counting Templates (QCTs)^28^ were designed for each amplicon. The predicted amplicon sequence for each primer pair was determined using Bowtie and the converted human genome, and 17 bases of flanking genomic sequence were added to both 5’ and 3’ ends. Eleven bases in the insert region of the QCT were replaced with Ns, allowing for a small number of unique QCTs to be added to each PCR reaction. QCTs were ordered from either Eurofins Genomics (Louisville, KY) and IDT (Coralville, IA). QCTs were diluted to 200 molecules per PCR reaction at each amplicon.

### Specimen sourcing and processing

To obtain a number of tumor specimens spanning a variety of tissue types, banked flash frozen tumors and buffy coats from the same subjects were obtained from Spectrum Health (Grand Rapids, MI).

To compare the performance of Northstar Response with an orthogonal treatment select based assay, banked plasma and matched buffy coat from metastatic lung cancer patients were processed through both assays. Collection of patient samples and subsequent genomic studies were approved by the University of California San Diego Institutional Review Board, and patients provided written informed consent to participating in the study prior to enrollment. Whole blood was collected in K2 EDTA tubes and processed immediately or within 2 hours after storage at 4C. Plasma and cellular components were separated by centrifugation at 800 g for 10 minutes at 4C. Plasma was centrifuged a second time at 18,000 g at room temperature to remove any remaining cellular debris. Plasma and buffy coat samples were stored at 80C until the time of DNA extraction.

To test whether the assay could detect changes in methylation that were concordant with clinical outcomes, samples were prospectively collected from cancer patients through contract research organizations (iSpecimen, Accio Biobank Online) and their partner clinics. Each clinical study was performed under the approval of their respective institution’s Institutional Review Board, and patients provided written informed consent to participate in the study prior to enrollment. Patients that were diagnosed with cancer and had not started treatment were enrolled. Up to 25 mL of blood was collected in Streck tubes (La Vista, NE) pre-treatment and at subsequent time points post-treatment. Blood tubes were then shipped from the clinic to BillionToOne’s laboratory. Clinical outcomes were provided when available.

Plasma and buffy coat were isolated from Streck tubes within 3 days of collection. Plasma and cellular components were separated by centrifugation at 1,600 g for 10 minutes at 22C. Plasma was centrifuged a second time at 16,000 g for 10 minutes without temperature control to remove any remaining cellular debris. cfDNA was extracted using the QIAamp Circulating Nucleic Acid Kit (Qiagen, Hilden, Germany), and gDNA was extracted from tumor samples and buffy coat samples using the DNeasy Blood & Tissue Kit (Qiagen, Hilden, Germany).

Contrived samples resembling cfDNA from cancer patients were created by mixing either 30 ng total of tumor gDNA and buffy coat gDNA at various tumor fractions. To mimic the fragment length of cfDNA, all tumor and buffy coat gDNA samples were sheared with a Covaris E220 Sonicator (Covaris, Woburn, MA) to a fragment length of ∼175 bp prior to mixing. Replicates of contrived tumor samples from a stage IV lung adenocarcinoma and a stage IV breast adenocarcinoma were prepared by combining sheared tumor and paired sheared buffy coat DNA samples at 0%, 0.25%, 0.5%, 1%, and 2% tumor fraction by mass, assuming that the tumor was 100% cancerous tissue (N = 12 0% breast replicates; N = 13 0% lung replicates; N = 15 0.5% breast replicates; N = 16 for all other conditions). To prepare samples at 1% and 2% tumor fraction for tumors from a variety of tissue types, sheared tumor and buffy coat DNA samples were combined in different ratios to account for tumor purity. For instance, if we estimated a tumor-extracted DNA sample to be 20% pure, the 1% tumor fraction sample would have contained 5% tumor-extracted DNA to account for purity.

Analytical validation samples were bisulfite converted using the EZ-96 DNA Methylation-Lightning MagPrep kit (Zymo, Irvine, CA) for all analytical experiments, and clinical samples were bisulfite converted with the Premium Bisulfite kit (Diagenode, Denville, NJ) which uses identical incubation times and temperatures. When either cfDNA or gDNA sample volumes were larger than the recommended input volume, samples were split in half, converted separately, and then re-combined during purification steps before elution of the converted DNA.

Multiplex PCR was performed on bisulfite converted specimens using Q5U polymerase (NEB, Ipswich, MA). Subsequently, indexing PCR was performed using Q5 polymerase (NEB, Ipswich, MA) in order to sequence multiple samples on the same sequencing run with dual indexes. Pooled libraries were bead cleaned and loaded on NextSeq 2000 (Illumina, San Diego, CA) sequencing instruments using P3 100 cycle reagents for single-directional sequencing.

### Northstar Response bioinformatic processing

Fastq files were adapter trimmed on the 3’ end using BBDuk and then mapped using BWA-MEM to a custom genome composed of the target hypermethylated and highly methylated amplicon and QCT sequences. For each amplicon, reads that mapped to the target amplicon were binned based on sequence. The number of CpGs contained in each sequence was calculated, and each sequence was classified as belonging to a methylated read if the number of CpGs was greater than or equal to the maximum number of possible CpGs for that amplicon minus one. Reads mapping to the corresponding QCT sequence were separately processed and binned based on the random N sequence of the QCT. Assuming each QCT molecule’s sequence is unique in each reaction, an average number of reads per QCT molecule was calculated. The total number of methylated molecules for that amplicon can then be calculated by dividing the total number of methylated reads by the average reads per QCT molecule.

When measurements are obtained for paired cfDNA and buffy coat samples, one can reduce background levels of methylation by subtracting the buffy coat methylation signal from the cfDNA methylation signal on a per-locus basis. It is estimated that about 55% of the cfDNA is of white blood cell origin based on methylation patterns^31^, suggesting that subtracting the entirety of the buffy coat methylation is a conservative approach to remove background methylation signal. Any calculated negative numbers of molecules after background subtraction were capped at zero.

In order to perform a comparable subtraction between buffy coat and cfDNA samples, the samples must first be normalized to the input number of genomic equivalents. The average of the highly methylated loci methylated molecules is used to estimate the total number of genomic equivalents in the sample overall. The number of methylated molecules at each locus is normalized by the estimated genomic equivalents of that sample. The number of normalized methylated molecules is then used for background subtraction.

Based on testing healthy subjects during assay development, certain hypermethylation loci were found to have significant methylation signal even after buffy coat subtraction. To minimize the amount of false cancer signal, any loci found to either consistently contribute a moderate amount of methylation or occasionally contribute a high amount of methylation in healthy subjects were filtered out from analysis. These loci often contain high amounts of methylation in buffy coat. After filtering out noisy hypermethylation loci, the resulting Tumor Methylation Scores were used to establish a noise floor, below which the methylation signal is not interpretable because it is comparable to the amount seen in healthy subjects.

### Making calls

When two time points of data are available, a call can be made as to whether there has been an increase, decrease, or no change in the Tumor Methylation Score. The call is made by modeling each time point’s measurement as a normal distribution with a mean equal to the number of measured methylated molecules normalized to the total number of input molecules. A standard deviation is assigned to the normal distribution based on the total number of methylated molecules. To assign these standard deviations, we use our prior measurements of the coefficient of variation at different tumor fractions for two tumors (Figure 2A). Specifically, we used this data to build a logarithmic regression model for the coefficient of variation as a function of Tumor Methylation Score, accounting for expected Poisson sampling noise. We utilize this model to extrapolate the coefficient of variation for any given distribution, then compute the standard deviation as the product of the coefficient of variation and distribution mean. The normal distribution from the first time point is subtracted from the second time point and normalized to the mean of the first time point’s normal distribution, creating a normalized difference normal distribution. An Increase call is made if the mean of this distribution is ≥15% and the log2 likelihood ratio that the difference is ≥15% compared to <15% is greater than 3. Similarly, a Decrease call is made if the mean of this distribution is ≤-15% and the log2 likelihood ratio that the difference is ≤-15% compared to >-15% is greater than 3. If the relative difference is not of sufficiently large magnitude or the statistical likelihood is not sufficiently strong, a No Change call is made. If either time point is below the noise floor, the mean for that time point is set to the noise floor, the standard deviation is still calculated based on the total number of methylated molecules, and a call can still be made. If the methylation signal from both time points is below the noise floor, an Indeterminate call is made.

### *In Silico* determination of Tumor Methylation Scores at untested tumor fractions

To analyze the assay’s performance for step increases of 0.25% and 0.5% tumor fraction, we simulated Tumor Methylation Scores at tumor fractions that were not experimentally tested. For simulated tumor fractions between 0% and 1%, the measured Tumor Methylation Scores of 1% tumor fraction replicates were multiplied by the ratio of the desired tumor fraction and the original tumor fraction to yield simulated Tumor Methylation Scores. Similarly, Tumor Methylation Scores for simulated tumor fractions between 1% and 2% were calculated by appropriately scaling down the Tumor Methylation Scores of 2% tumor fraction replicates.

To determine the sensitivity of the assay at low tumor fractions across multiple tissue types, we employed a similar approach to simulate Tumor Methylation Scores at lower tumor fractions. Tumor Methylation Scores measured at 1% and 2% tumor fraction were each multiplied by the same ratio to simulate smaller absolute changes in tumor fraction that still reflect a doubling.

### VAF-based somatic mutation assay

An in-house hybrid capture-based assay was used to measure VAF of somatic mutations. The assay consists of the addition of UMIs and universal amplification of samples using an IDT xGen™ cfDNA & FFPE DNA Library Preparation Kit and target enrichment using an IDT xGen™ Custom Hybridization Capture Panel (IDT, Coralville, IA). The target enriched libraries were run on a NextSeq 2000 sequencer (Illumina, San Diego, CA) at approximately 100 million reads per sample. Mutations were called using a custom-built bioinformatics pipeline, which involves UMI consensus calling of molecules, filtering and thresholding of data to reduce background and erroneous signals, and calling of somatic mutations with VAF determined based on the ratio of mutant to wild type molecule counts. The clinical significance of any detected mutations was assessed using Qiagen Clinical Insight (Qiagen, Hilden, Germany). Only clinically significant mutations were included for VAF analysis.

To determine the performance of VAF-based therapy response monitoring approaches, samples were prepared by extracting genomic DNA from 10 different tumor samples with known mutations, and one healthy tissue sample. These genomic DNA samples were sheared via sonication with a Covaris E220 Sonicator (Covaris, Woburn, MA) to an average length of ∼175 bp to mimic the size of native cfDNA fragments. To generate a wide range of mutation VAFs in the final sample, the tumor samples were combined at equal proportions to target various allele frequencies, and the tumor mixture was sequenced in 6 replicates to determine the VAF in the mix for each variant. The combined tumor mix sheared DNA sample was added into a healthy background (buffy coat sheared DNA) sample at 10% and 20% by mass. 20 replicates of 30 ng of each of these mixes were run on an in-house hybrid capture-based assay. Mutations were filtered by VAF to create groups of mutations centered around each VAF of interest: 0.25% (8 mutations ranging from 0.2% to 0.3%, with mean 0.25%), 0.5% (10 mutations ranging from 0.4% to 0.6%, with mean 0.52%), 1% (10 mutations ranging from 0.8% to 1.2%, with mean 0.97%), and 2% (5 mutations ranging from 1.2% to 2.6%, with mean 1.77%).

In order to determine whether the performance of the VAF-based assay is representative of the performance of VAF-based approaches in general, a simulation was performed to determine whether the noise in the VAF measurement agrees with statistical theory. Data was generated assuming Poisson sampling of mutant molecules around a mean of the number of mutant molecules (central VAF [0.25%, 0.5%, 1%, 2%) x 2000 genomic equivalents (g.e.)], with 10000 mutations simulated for each VAF. 2000 g.e. was chosen because it was the average detected across all the mutations in the real VAF dataset. The data was split into subgroups of different numbers of mutations and the VAF was summed across them, then the noise was calculated by taking the CV for each individual or summed VAF for each group size. The simulation was rerun 100 times to determine the variability in this measurement. The median across the 100 replicates of the simulation is shown in Figure 4, for 1, 2, 3, and 4 mutations utilized. The full data is shown in Supplementary Figure 3.

### Methylation-based estimation of tumor purity

One challenge that complicates comparisons of assay performance across many different tumors is the unknown fraction of cancerous tissue in each specimen. To account for this, we tested matched tumor and buffy gDNA for each subject and calculated a tumor purity score for each tumor. Tumor purity scores were calculated as follows: 1) The number of methylated molecules detected at each hypermethylation locus was normalized to 1000 genomic equivalents, as measured by highly methylated control loci; 2) The percent hypermethylation was calculated for each locus as the difference between the number of methylated molecules measured in tumor gDNA and buffy coat gDNA divided by 1000 minus the number of methylated molecules measured in buffy coat; 3) The tumor purity of a given tumor was defined as the 75th percentile of percent hypermethylation across all informative loci detected in a tumor sample. The tumor purity scores ranged from 0.7% to 104% across tumors, and we capped the estimated tumor purity at 100% when accounting for tumor purity to make contrived samples at 1% and 2% tumor fraction (Supplementary Figure 4A).

To validate the computed tumor purity score as a meaningful metric, we compared tumor purity scores for a variety of tumors to orthogonal tumor purity estimates that were based on somatic mutations. We used a least squares approach to identify a percentile that maximizes concordance between the two orthogonal measurements of tumor purity, which is how we decided to define the tumor purity as the 75th percentile of percent hypermethylation across loci (Supplementary Figure 4B, 4C).

## Data Availability

The ability to share primary sequence data was not included in the informed consent signed by subjects. A data use agreement is required to access the data.

## Acknowledgements

We would like to acknowledge Naomi Searle for her expertise using QCI for variant classification. We would like to acknowledge Matthew Varga, John ten Bosch, and Shan Riku for providing editorial support.

## Author Contributions

PY, RV, KS, XB, WZ, HH, DT, and OA conceived and designed experiments. RV, KS, SL, XB, JW, JZ, and BW conducted experiments. PY, RV, KS, XB, JW, and WZ analyzed the results. PY, RV, KS, and XB wrote the manuscript. All authors reviewed the manuscript.

---

**Supplementary Figure 1.**
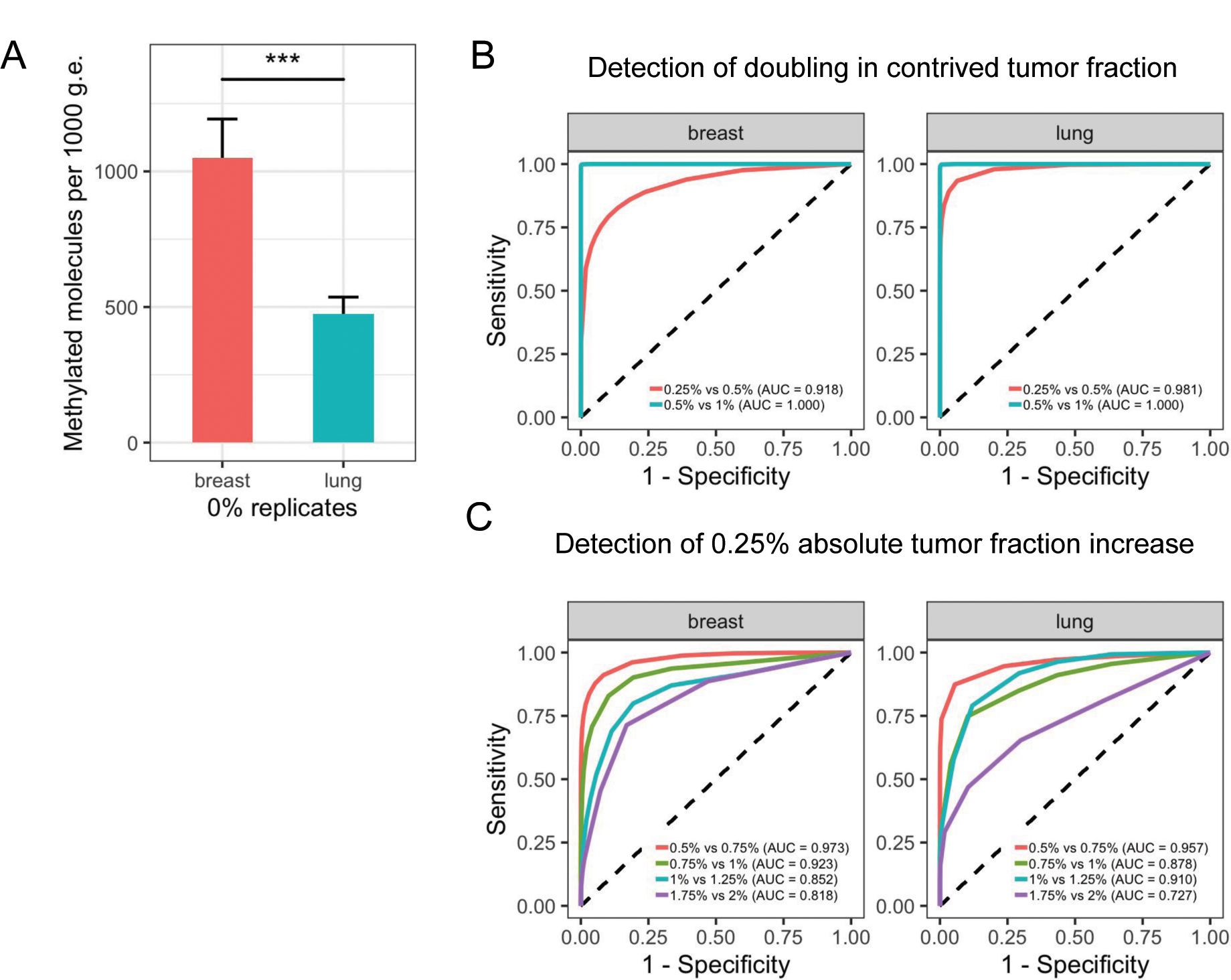
(**A**) Total number of methylated molecules in 0% tumor replicates, normalized to 1000 input g.e. for each sample. Error bars represent 2 standard deviations from the mean. *p* value = 1.2×10^-13^ (two-sided T test). (**B**) Receiver operating characteristic (ROC) curves for the sensitivity of calling changes in Tumor Methylation Score after a doubling of contrived tumor fraction. Specificity was measured for comparisons of the smaller tumor fraction against itself. (**C**) ROC curves for calling changes in Tumor Methylation Score after an increase of 0.25% contrived tumor fraction. Data for 0.75%, 1.25%, and 1.75% tumor fraction was obtained by appropriately scaling down the signal from 1% and 2% replicates, respectively.

**Supplementary Figure 2.**
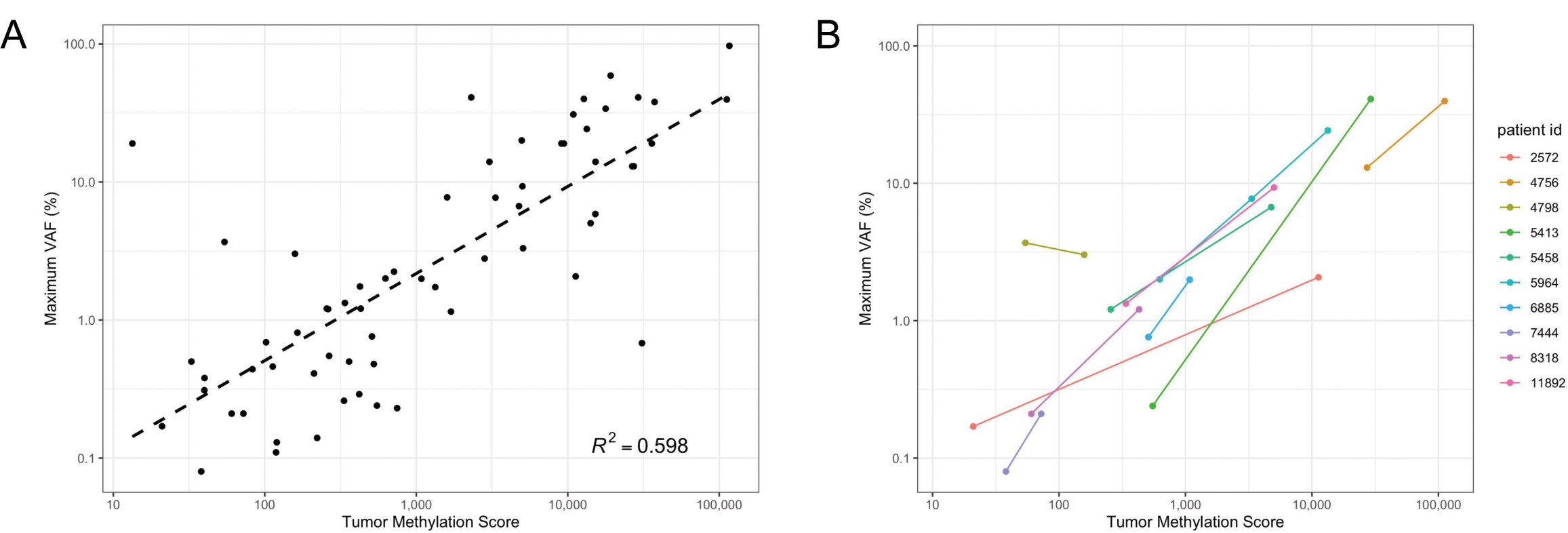
Correlation of Tumor Methylation Score with VAF in clinical samples without excluding clonal hematopoietic of indeterminate potential (CHIP) mutations. (**A**) Correlation of maximum VAF as measured by a treatment selection assay with Tumor Methylation Score in clinical samples (N = 64 samples). A linear fit and corresponding *R*^2^ was calculated using the logarithm of maximum VAF and the logarithm of Tumor Methylation Score. (**B**) Correlation of Tumor Methylation Score with maximum VAF within patients for therapy response monitoring (N = 10 patients).

**Supplementary Figure 3.**
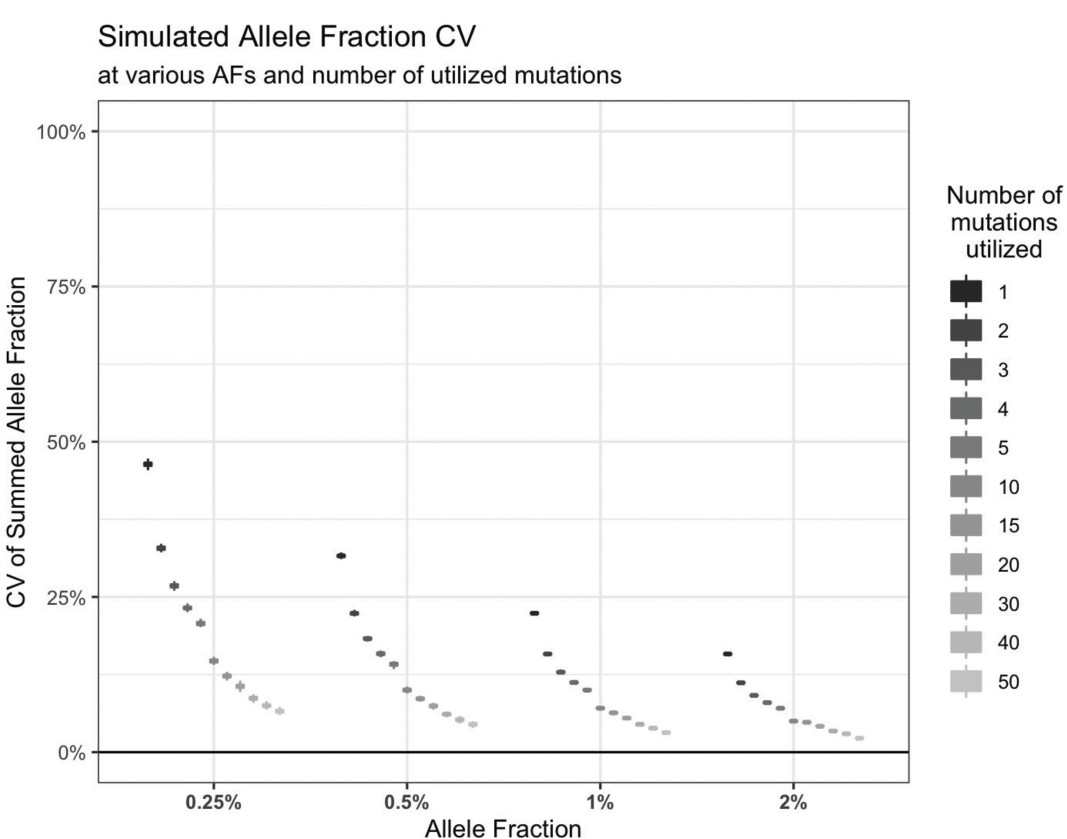
CV of summed variant allele fraction across multiple simulated Hybrid Capture mutations decreases both with increasing number of variants utilized and with increasing mutation variant allele fraction. Data were generated assuming Poisson sampling of mutant molecules around a mean of the number of mutant molecules (central VAF x 2000 g.e.), with 10000 mutations simulated for each VAF. All possible subgroupings of size N of the mutations at each VAF range were used to generate the range of CVs, where N is the number of mutations utilized. The medians for boxplots for 2, 3, and 4 mutations utilized are reproduced as stars in Figure 4.

**Supplementary Figure 4.**
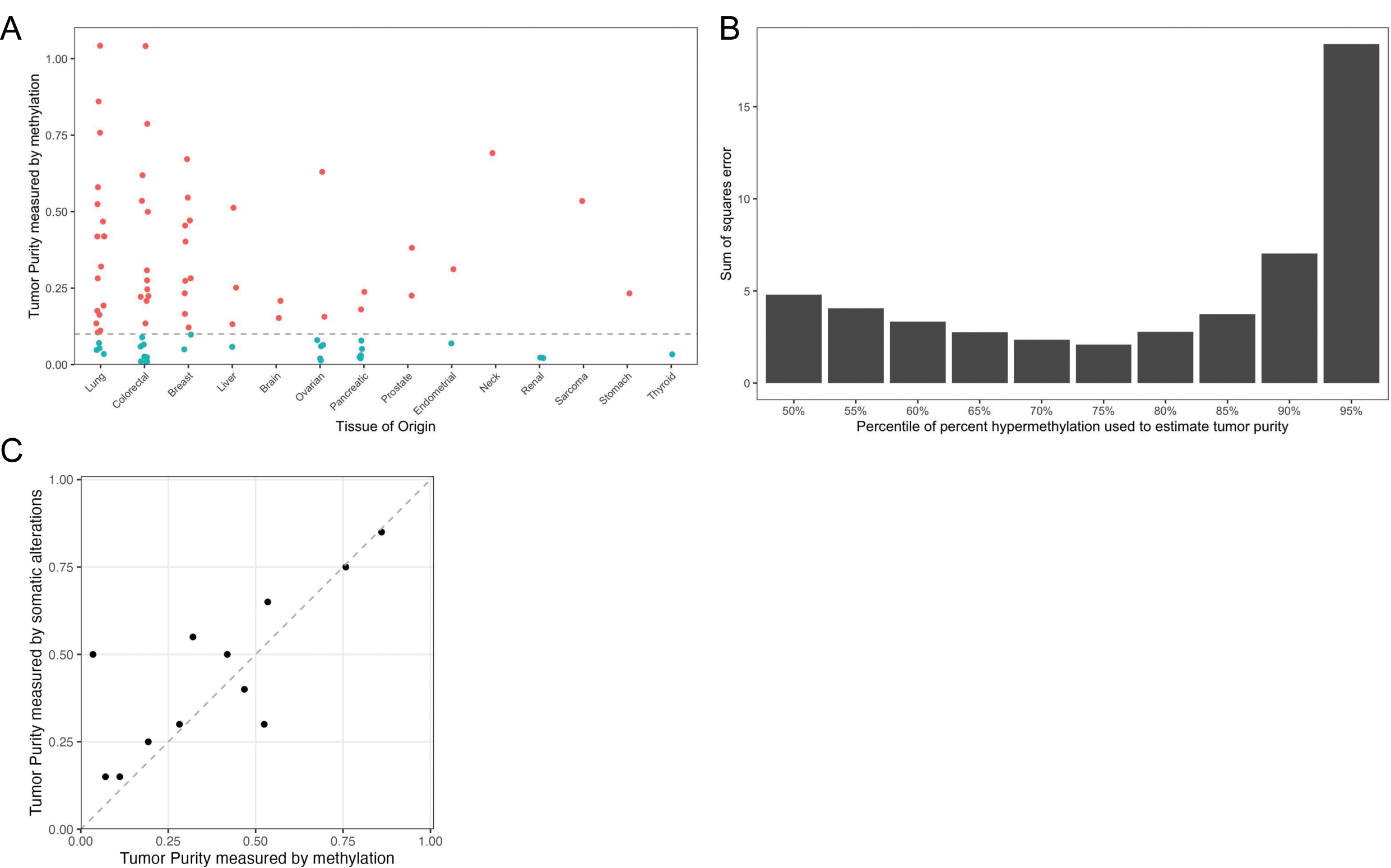
(**A**) Tumor purity scores based on methylation were calculated for 84 tumor specimens, and 54 tumors above 10% purity were selected for further testing. To calculate tumor purity scores, the number of methylated molecules at each hypermethylation locus was normalized to 1000 genomic equivalents as measured by highly methylated normalization loci. The percent hypermethylation of each locus was calculated as the difference between the number of methylated molecules measured in tumor gDNA and buffy coat gDNA divided by 1000 minus the number of methylated molecules measured in buffy coat. The 75th percentile of percent hypermethylation across all informative loci detected in a tumor sample was defined as the tumor purity. (**B**) The optimal percentile for estimating tumor purity was determined by testing different percentiles of percent hypermethylation and computing the sum of squares error with an orthogonal measurement of tumor purity based on somatic alterations including SNVs, indels, and copy number alterations. Using the 75th percentile of percent hypermethylation across all loci present in a tumor resulted in the lowest sum of squares error. (**C**) For a set of 12 lung tumors, the tumor purity measured by methylation using the 75th percentile method correlates with tumor purity measurements based on somatic alterations.

**Supplementary Figure 5.**
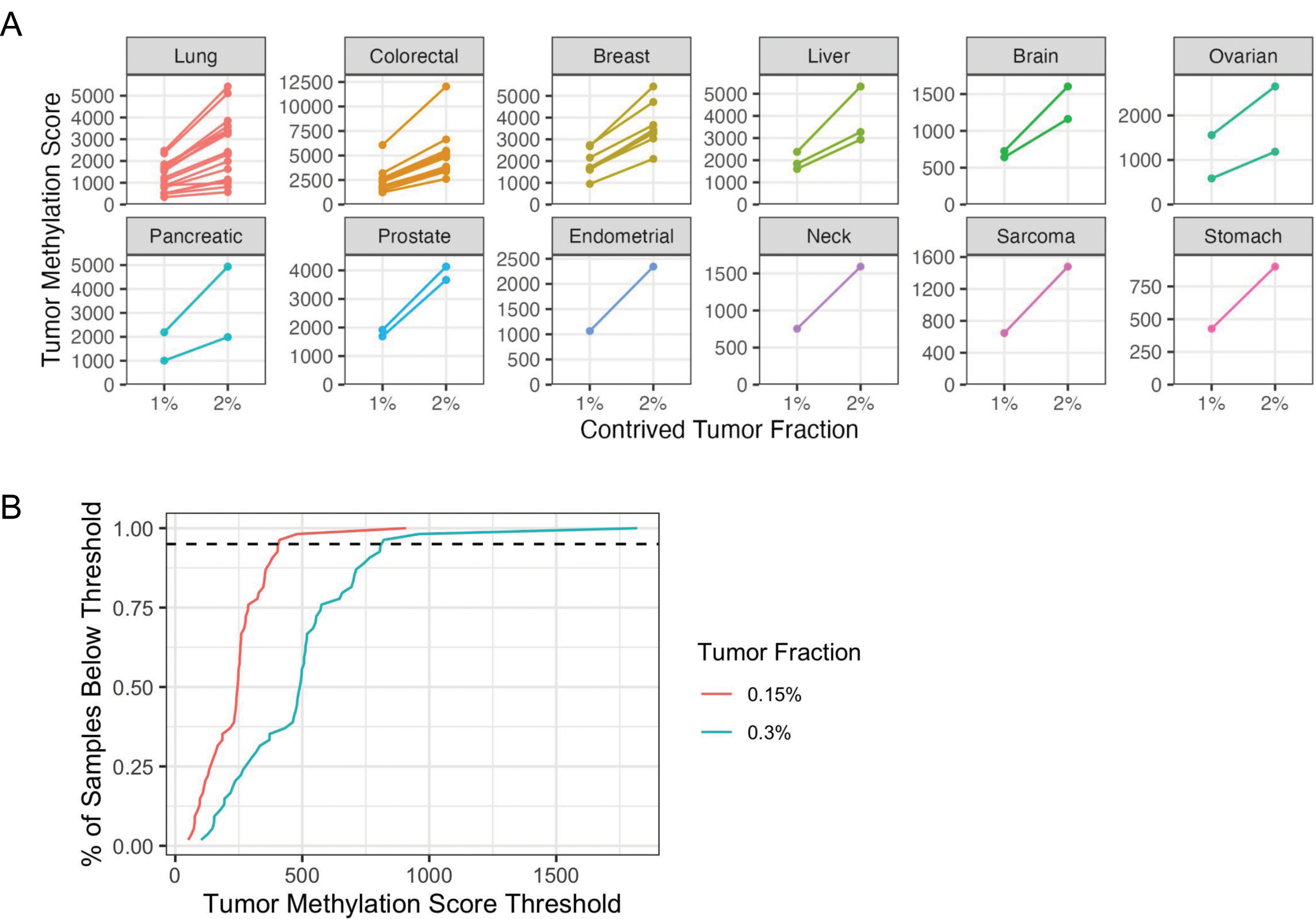
(A) Tumor Methylation Scores detected at 1% and 2% contrived tumor fraction for 54 different tumor specimens. **(B)** Cumulative distribution of Tumor Methylation Score across simulated contrived tumor samples. Tumor Methylation Scores at 0.15% and 0.3% were obtained by linearly scaling down the signal of 1% contrived tumor samples.

**Supplementary Figure 6.**
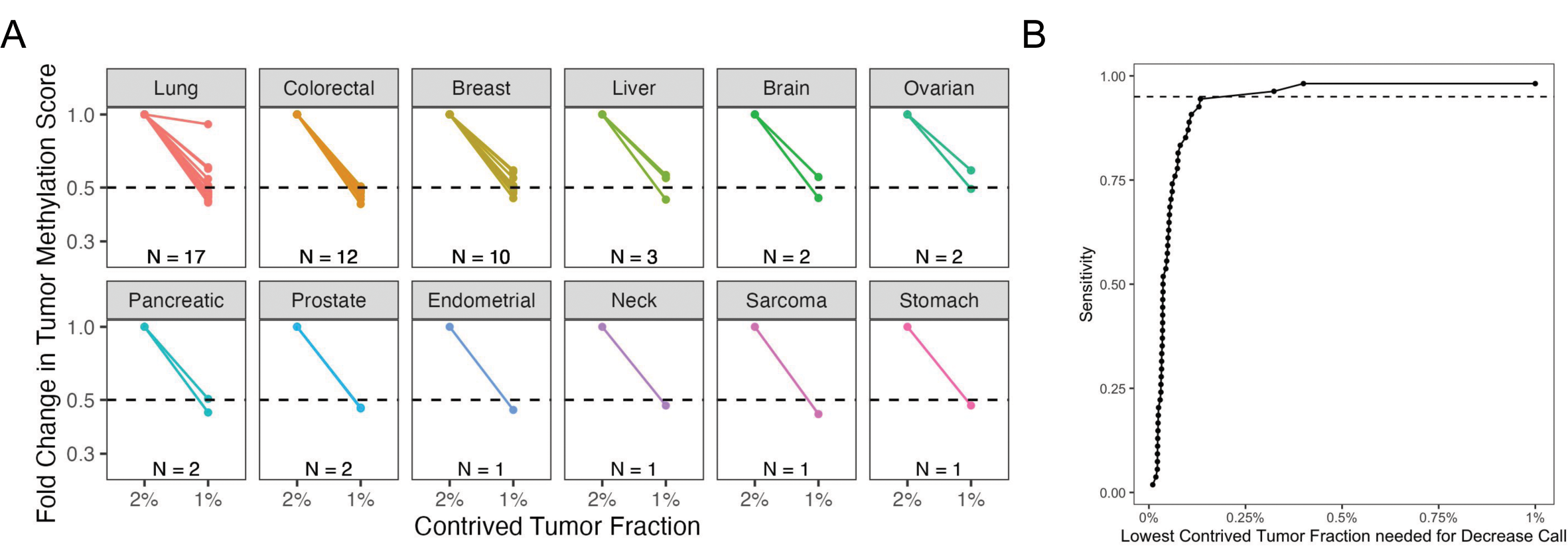
(**A**) When we made comparisons between contrived samples to reflect a halving in tumor fraction rather than a doubling, Tumor Methylation Scores were significantly decreased at 1% tumor fraction relative to 2% tumor fraction for 53/54 different tumor specimens. (**B**) The lowest contrived tumor fractions at which decrease calls could have been made were estimated by linearly scaling down the Tumor Methylation Scores at 1% and 2% tumor fraction *in silico*, maintaining the same fold-difference in Tumor Methylation Score. At 0.32% tumor fraction, decrease calls could still be made for 52/54 tumor specimens, which corresponds to 96.3% sensitivity.

**Supplementary Figure 7.**
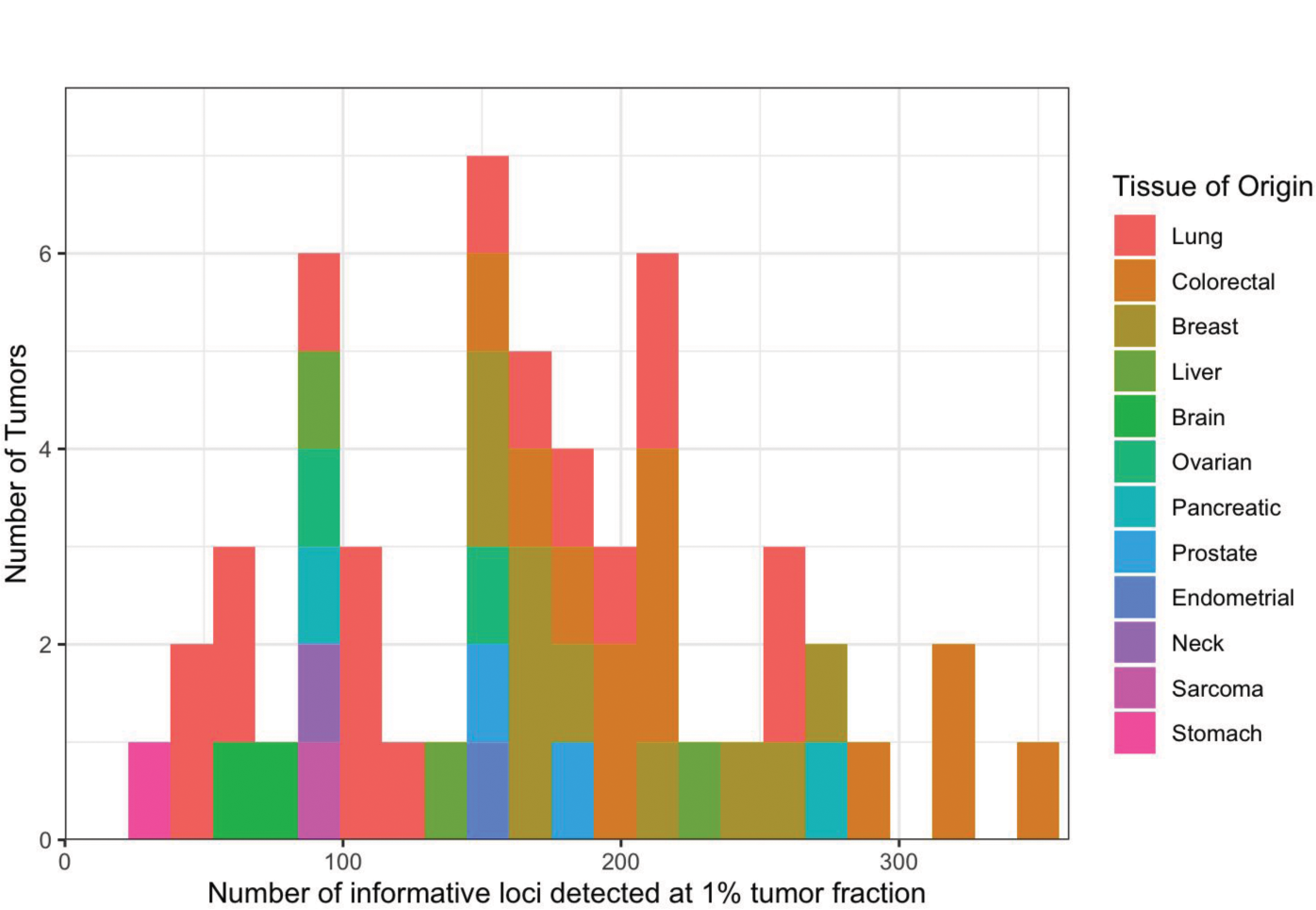
Across a set of 54 tumors, the number of informative hypermethylation loci detected at 1% contrived tumor fraction ranged from a minimum of 28 to a maximum of 350 loci.

**Supplementary Figure 8.**
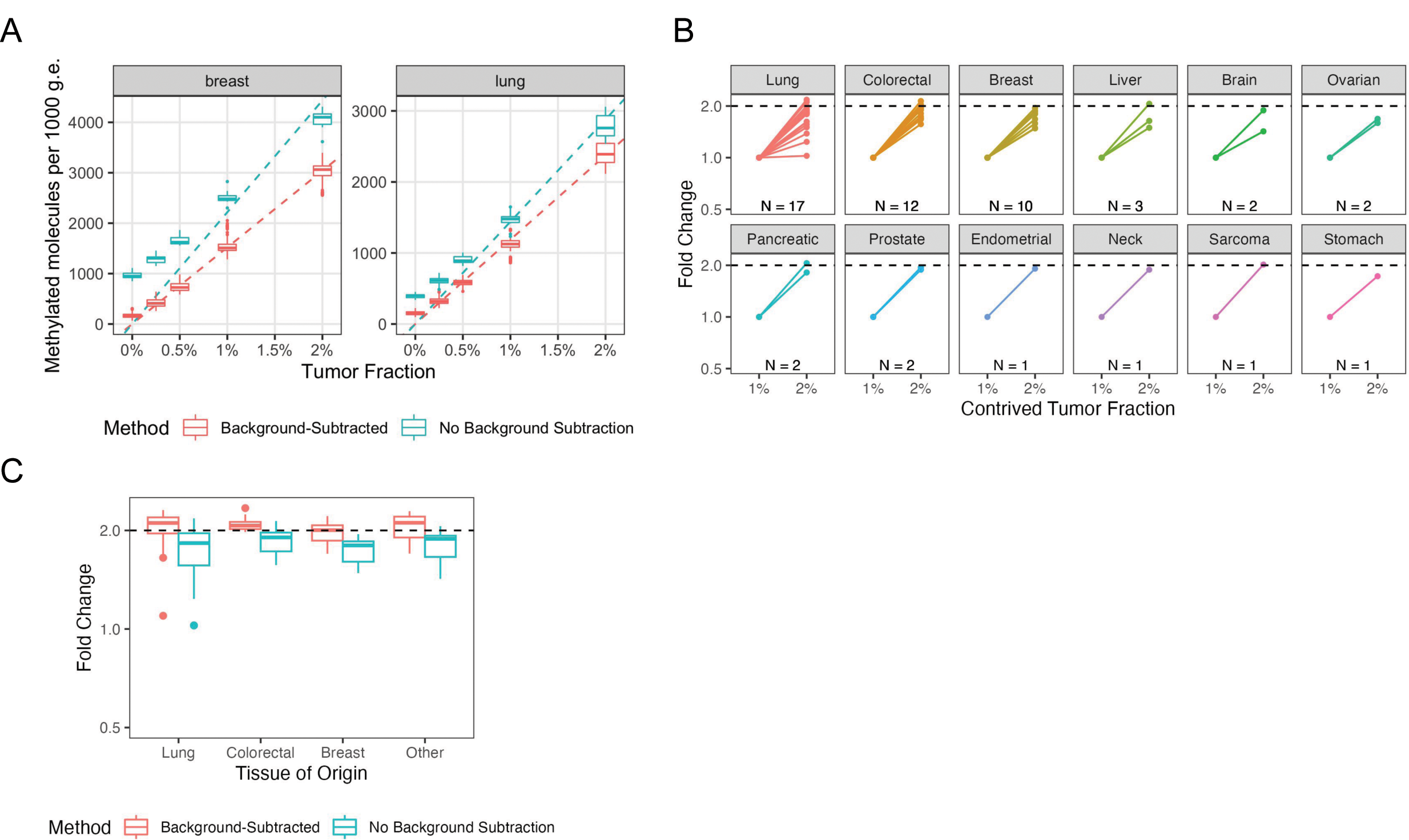
Utility of background subtraction for calculating Tumor Methylation Score. (A) The number of methylated molecules, normalized to 1000 g.e. input, was measured in technical replicates of contrived samples at different tumor fractions with and without background subtraction of methylated molecules detected in the paired buffy coat. The dashed lines indicate a linear regression *y = mx* through the mean number of methylated molecules at each tumor fraction (background-subtracted: breast *R*^2^ = 0.998, lung *R*^2^ = 0.996; no background subtraction: breast *R*^2^ = 0.929, lung *R*^2^ = 0.977). (B) The fold change in the number of methylated molecules detected per 1000 g.e. without removing background signal detected in buffy coat. (C) When there is a doubling in contrived tumor fraction, the fold change in the number of methylated molecules detected per 1000 g.e. is more accurate and precise when a background subtraction is performed.

